# Performance evaluation of virus concentration methods for implementing SARS-CoV-2 wastewater based epidemiology emphasizing quick data turnaround

**DOI:** 10.1101/2021.05.29.21257950

**Authors:** Md Ariful Islam Juel, Nicholas Stark, Bridgette Nicolosi, Jordan Lontai, Kevin Lambirth, Jessica Schlueter, Cynthia Gibas, Mariya Munir

**Affiliations:** Department of Civil and Environmental Engineering, 9201 University City Blvd, Charlotte, NC, 28223; Department of Bioinformatics and Genomics, 9201 University City Blvd, Charlotte, NC, 28223; Bioinformatics Research Center, 9201 University City Blvd, Charlotte, NC, 28223; Department of Leather Engineering, Khulna University of Engineering and Technology, Khulna, 9203

**Keywords:** SARS-CoV-2, Innovaprep CP Select, Electronegative Membrane Filtration, Virus concentration, Wastewater based epidemiology (WBE)

## Abstract

Wastewater based epidemiology (WBE) has drawn significant attention as an early warning tool to detect and predict the trajectory of COVID-19 cases in a community, in conjunction with public health data. This means of monitoring for outbreaks has been used at municipal wastewater treatment centers to analyze COVID-19 trends in entire communities, as well as by universities and other community living environments to monitor COVID-19 spread in buildings. Sample concentration is crucial, especially when viral abundance in raw wastewater is below the threshold of detection by RT-qPCR analysis. We evaluated the performance of a rapid ultrafiltration-based virus concentration method using InnovaPrep Concentrating Pipette (CP) Select and compared this to the established electronegative membrane filtration (EMF) method. We evaluated sensitivity of SARS-CoV-2 quantification, surrogate virus recovery rate, and sample processing time. Results suggest that the CP Select concentrator is more efficient at concentrating SARS-CoV-2 from wastewater compared to the EMF method. About 25% of samples that tested negative when concentrated with the EMF method produced a positive signal with the CP Select protocol. Increased recovery of the surrogate virus control using the CP Select confirms this observation. We optimized the CP Select protocol by adding AVL lysis buffer and sonication, to increase the recovery of virus. Sonication increased Bovine Coronavirus (BCoV) recovery by 19%, which seems to compensate for viral loss during centrifugation. Filtration time decreases by approximately 30% when using the CP Select protocol, making this an optimal choice for building surveillance applications where quick turnaround time is necessary.

## 1. Introduction

Wastewater-based epidemiology (WBE) is a widely used approach that has been rapidly adopted by the environmental science and engineering academic community as part of the response to the COVID-19 pandemic. WBE has been demonstrated to be an effective early warning tool for rising case numbers, when combining COVID-19 wastewater surveillance data and public health data. As it can provide evidence of both symptomatic and asymptomatic COVID-19 cases, WBE has been applied to detect COVID-19 cases in college residence halls (Betancourt et al., 2021; Gibas et al., 2021; Harris-Lovett et al., 2021; Scott et al., 2021), schools (Gutierrez et al., 2021; Crowe et al., 2021), nursing homes (Spurbeck et al., 2021), and other group living settings. Precise and accurate quantification of viral copies in wastewater is a prerequisite for a successful WBE surveillance project. Detection sensitivity is dependent on the choice of an effective and reliable virus concentration method prior to RNA extraction and quantification.

Virus concentration is crucial in the wastewater especially when viral titers are very low, as is seen in building-based surveillance (Corchis-Scott et al., 2021; Gibas et al., 2021). Polyethylene glycol (PEG)-based precipitation was initially widely used to concentrate the virus with successful signal detection ((La Rosa et al., 2020; Wu et al., 2020a; Kumar et al., 2020). This method, however, requires a long processing time. Other methods such as Electronegative Membrane Filtration (EMF) which also known as HA method, and Ultrafiltration have been used successfully to concentrate viruses from wastewater prior to RNA extraction in a variety of application contexts worldwide. (Ahmed et al., 2020a; Medema et al., 2020; Nemudryi et al., 2020; Wu et al., 2020b; Wurtzer et al., 2020). Skimmed milk flocculation is suggested as a promising method for resource limited environments based on its detection consistency and simplicity (Philo et al., 2021). Another study focused on a two-step concentration procedure to process large wastewater volumes (McMinn et al., 2021). Among the available methods, the EMF method has previously been reported to be one of the most efficient methods of virus concentration based on surrogate virus recovery rate (Ahmed et al., 2020a). However, Jafferali et al., (2021) recently reported that ultracentrifuge-based methods showed better efficiency in spike recovery and quantification of SARS-CoV-2, citing qPCR inhibition as a potential pitfall of the EMF method.

We previously reported outcomes of building level surveillance WBE for a large urban college campus during Fall 2020 using EMF as the method of concentration (Gibas et al., 2021). However, to shorten the timeline from sample collection to reporting, we have tested and adopted an alternative concentration method using the InnovaPrep CP Select rapid concentrator. The CP Select is an automatic system that allows the user to concentrate bacteria or virus particles by passing a liquid sample through either hollow or ultrafiltration based concentrating pipette tips. It can process large volumes (up to 5 L) depending on the turbidity of the sample and can concentrate to volumes as small as 150 uL (https://www.innovaprep.com). Rusiñol et al., (2020) investigated three rapid concentration methods: skimmed milk flocculation (SMF), InnovaPrep CP Select automated ultrafiltration using (150 kDa) filter tips, and centrifugal-ultrafiltration (CeUF) using the Centricon plus-70 (100 kDa) using MS2 as the surrogate virus spiked into wastewater samples. The higher MS2 recovery (51%) in this study was achieved using the InnovaPrep quick concentrating pipette (CP) after removing debris compared to SMF (29%) and CeUF (16.5%). Limited replication in that study did not allow for a firm conclusion, and the use of MS2, a non-enveloped virus, as a surrogate was not optimal as a benchmark for recovery of an enveloped virus like SARS-CoV-2. Gonzalez et al., (2020) reported the use of the CP Select concentrator for COVID-19 surveillance in the southeastern Virginia area, and performed a comparison of viral surrogates from treatment plant influent wastewater, in which the CP Select reported an average BCoV recovery of 5.5% compared to 4.8% with EMF. The characteristics of wastewater collected from congregate living facilities such as university residence halls, schools, and nursing homes are somewhat different from the highly pooled wastewater treatment plant (WWTP) influent. Building level wastewater has a higher variability in viral load, fecal matter content and suspended solids concentration compared to WWTP influent samples. Corchis-Scott et al., (2021) reported that the pepper mild mottle virus (PMMoV), a fecal indicator, in residence hall wastewater varied in concentration across 4 orders of magnitude with a coefficient of variation (CV) of 2.83. In comparison, the signal varied more modestly for influent samples from five different WWTPs, with concentrations falling within only one order of magnitude (CV of 0.38). Because our surveillance system relies on raw building-level wastewater, we have evaluated the CP Select specifically in the building surveillance context with a direct comparison to the established EMF method. We aimed to determine how the optimized CP Select method performs compared to the EMF method in terms of filtration time, BCoV recovery, and sensitivity of SARS-CoV-2 detection and quantification.

We also investigated whether, due to the complex nature of the wastewater, RNA extracted from wastewater might contain inhibitors to RT-qPCR amplification, by evaluating inhibition under each concentration protocol. These optimizations resulted in a CP Select protocol providing increased viral recovery and suitable for rapid reporting of results from building-level SARS-CoV-2 WBE.

## 2. Experimental Method

### 2.1 Sample collection

In conjunction with the COVID-19 Wastewater Surveillance being conducted on the UNC Charlotte campus (Gibas et al., 2021), we collected samples from thirty-seven sites that were used to monitor a combination of dormitories, greek village housing and neighborhood sites consisting of on-campus non-residential buildings. Wastewater samples were collected thrice weekly via HACH AS960 and ISCO GLS Compact autosampler devices located at a building plumbing cleanout or at a manhole accessed externally. At each of these sites, an autosampler was placed on flat ground at higher elevation than the sample stream. The autosampler was connected via a ⅜” tubing coupler to HACH silicone rubber pump tubing. At cleanout sites, ISCO Silicone rubber pump tubing was directly connected to a double-sided tubing connector on a cap externally located on the plumbing clean out. Additional tubing was connected to the underside of the cap to reach the sample stream. At manhole sites, tubing was fed through a cut out in the manhole lid, and then through a PVC guide to the sample stream. The end of the tubing that resides in the sample stream was bound to a strainer, designed to filter out large solids and prevent build up in suction lines. A drawing of the autosampler setup for manhole and cleanout cap location of the sewage pipeline is available in our previously published paper (Gibas et al., 2021). Each autosampler is powered by a 12V lead acid battery and contains a 9.46 L Nalgene sample bottle in the HACH AS960 devices, or a 3500 mL sample bottle in the ISCO Compact devices. Autosampler devices were similarly programmed to draw ∼20 mL of wastewater every 30 minutes for 24 hours, to generate a composite sample for lab processing. Upon collection, each composite sample was divided into several 50 mL centrifuge tubes to be used in this experiment, for routine surveillance testing, or for archiving. A total of 53 wastewater samples were collected during five separate sampling events between October 2020 and March 2021for this study.

### 2.2 Sample volume processing/filtration threshold

Ten samples were used to test the impact of turbidity on sample processing time (Table 1). VWR/BDH Chemicals pH test strips and the HACH 2100Q Portable Turbidimeter were used to determine pH and turbidity, respectively. The maximum value that can be accurately determined using the HACH 2100Q Portable Turbidimeter is 1000 NTU. Any value that exceeds this limit was listed as >1000 NTU. EMF (HA filtration) was routinely used as the virus concentration method for SARS-CoV-2 surveillance as previously reported (Gibas et al., 2021). When 40-50 mL wastewater samples were processed, turbid samples require a long processing time, due to clogging of filter pores. In preliminary tests, the InnovaPrep CP Select concentrator was capable of processing 125-150 mL wastewater samples, regardless of turbidity. We compared the filtration capability of both EMF and the CP Select protocols systematically, by processing 40 - 100 mL volumes of 10 different samples using each method. We chose 5 samples which were turbid and 5 which were visually clear, excluding samples that exceeded the measurement threshold for turbidity. Processing time was recorded for each input volume, and downstream outcomes in viral surrogate recovery as well as in the qPCR detection step were compared.

**Table 1.** Filtering volume time comparison between EMF and CP Select method.

| Sample ID | pH | Turbidity (NTU) | 40 mL filtering (min) |  | 60 mL filtering (min) |  | 100 mL filtering (min) |  |
| --- | --- | --- | --- | --- | --- | --- | --- | --- |
|  |  |  | EMF | CP Select | EMF | CP Select | EMF | CP Select |
| S1 | 7 | 26.7 | 1 | 5 | 2 | 10 | Over 30 | 20 |
| S2 | 7 | 27.3 | 1 | 1 | 2 | 1 | Over 30 | 5 |
| S3 | 7.5 | 13.2 | 1 | 1 | 1 | 10 | 6 | 10 |
| S4 | 9 | 53 | 1 | 2 | Over 30 | 10 | Over 30 | 20 |
| S5 | 7.5 | 15.6 | 1 | 1 | 1 | 10 | 2 | 30 |
| S6 | 7 | 10.1 | 1 | 1 | 2 | 1 | 2 | 5 |
| S7 | 7.5 | 11 | - | 1 | - | 1 | - | 14 |
| S8 | 7 | 1000 | Over 30 | 2 | Over 30 | 7 | Over 30 | 30 |
| S9 | 8.5 | 347 | Over 30 | 1 | Over 30 | 10 | Over 30 | 20 |
| S10 | 7.5 | 97.8 | 7 | 2 | Over 30 | 10 | Over 30 | 14 |

### 2.3 Virus concentration and RNA Extraction

Bovine Coronavirus or BCoV (BOVILIS® Coronavirus, Merck Animal Health, NE, USA), a surrogate of human coronavirus, was spiked into the wastewater as a process control prior to sample concentration. The concentration of BCoV was previously determined (2.2×10^5 copies/mL) using ddPCR and spiked in at a concentration of 1uL per mL of wastewater. Samples were then processed via EMF filtration as previously described (Gibas et al., 2021). Briefly, wastewater samples were acidified to adjust the pH in the range of 3.5 - 4.0 followed by the addition of 100X MgCl_2_, 6H_2_0 (2.5M) in a ratio of 1:100 (Ahmed et al., 2020a; Ciesielski et al., 2021). 40 - 100 mL aliquots of adjusted wastewater were filtered through a 0.45 μm pore size, 47 mm diameter electronegative membrane filter (HA, Millipore) coupled with a disposable filter funnel (Pall corporation, NY, USA) until all liquid appeared to have passed through the filter. After filtration, the membrane filter was folded and resuspended in a 2 mL sterile tube containing 1000 uL of AVL lysis buffer with carrier RNA (Qiagen). The membrane filter suspended in the lysis buffer was incubated at room temperature for 10 minutes followed by vortexing for 15 sec to facilitate the recovery of adsorbed virus particles from the filter. For sample processing with the CP Select concentrator, wastewater samples were centrifuged for 10 mins at 1000011g to remove solid debris. 10% Tween-20 was added to the supernatant in a ratio of 1:100 before concentration, as recommended by the manufacturer to increase virus recovery. 40 to 150 mL samples were then filtered through a single use 0.05 µm PS Hollow Fiber Filter Tips (InnovaPrep) using the automatic CP Select™ (InnovaPrep). Viral particles attached to the filter tips were recovered by eluting with 0.075% Tween-20/Tris elution fluid using Wet Foam Elution™ technology (InnovaPrep) into a final volume ranging from 250 uL to 500 uL. Following the EMF or CP Select concentration step, we then used the QIAamp viral mini kit (Qiagen, Valencia, CA, USA) for RNA extraction from 200 uL of concentrated sample. RNA was extracted following the manufacturer-recommended protocol. Extracted RNA was eluted with AVE buffer into a final volume of 60 uL. All extracted RNA was stored at -80L until quantification.

### 2.4 RT-qPCR

Quantitative reverse transcription PCR (RT-qPCR) was used to detect and quantify SARS-CoV-2 and Bovine Coronavirus from extracted RNA. The CDC recommended N1 (Nucleocapsid) primer and probe set (Corman et al., 2020) was used for SARS-CoV-2 quantification while a primer/probe set published by Decaro et al., (2008) was used for Bovine Coronavirus quantification. All amplification reactions were carried out in one step, with a reaction volume of 20 µL. The SARS-CoV-2 assay consisted of 10 µL iTaq universal one step reaction mix (Bio-Rad, Hercules, CA), 0.5 µL iScript reverse transcriptase (Bio-Rad), 500 nM primers along with 125 nM probe (IDT), and 5.0 µL extracted RNA template. The reaction mix then was amplified using a CFX96 qPCR thermocycler (Bio-Rad, Hercules, CA) with the following thermocycling conditions: reverse transcription at 50°C for 15 min with initiation at 25°C for 2 minutes, followed by polymerase activation at 95°C for 2 min and 44 cycles of denaturation at 95°C for 3 s, followed by annealing at 55°C for 30 s (CDC, 2020). Single stranded RNA based SARS-CoV-2 positive control from Twist Bioscience was used to generate a standard curve using a series of ten-fold serial dilutions with concentrations ranging 10^5^ to 10 copies per reaction. All samples were run in triplicate along with a series of three positive and negative controls. The limit of detection (LoD) of assay was determined following the same protocol as described in Gibas et al., (2021). An extended dilution series of SARS-CoV-2 positive control in a range from 10^5^ to 1 copy/reaction in 6 replicates was amplified following the protocol for generating the standard curve as described above. The LoD of the RT-qPCR assay is determined as the lowest concentration at which all the replicates were positive with a less than 1 quantification cycle (Cq) variation among the replicates (Francy et al., 2012). The LoD of the assay was determined as 5 copies/reaction. The LoD of the method was then calculated by multiplying this concentration with the concentration factor which had been previously calculated considering the sample volume processed for the respective methods. Any samples to be considered as SARS-CoV-2 positive must have the concentration above the limit of detection with a minimum of two replicates agreement.

The BCoV assay was similar to the N1 assay, with the primer and probe concentrations at 600 nM and 200 nM, respectively. Thermal cycling parameters were the same used in the Decora et al. (2008) protocol, except the annealing temperature was set at 55°C instead of 60°C. This change improved the primer efficiency from 85% to 102.5%. For BCoV recovery quantification, a standard curve was generated using a serially diluted BCoV vaccine, in the concentration range of 10^5^ to 1 copies/reaction. All the primer and probe sequences and the standard curves are included in supplementary file (Table S5 and Figure S1, respectively). All samples were run in triplicate along with a series of three positive and negative controls. The BCoV recovery efficiency was calculated based on the following equation:

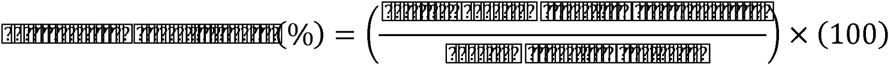

The effective volume assayed is the original volume of wastewater assayed prior to concentration per reaction in the RT-qPCR. This volume was determined considering initial wastewater volume used in the concentration step, the portion of the concentrated sample used in the RNA extraction process, and the volume (uL) of RNA used in the RT-qPCR reaction. While the concentration factor was calculated considering the initial volume of wastewater filtered and final elution volume after concentration step.

### 2.5 CP Select protocol optimization

The addition of AVL lysis buffer with carrier RNA (Qiagen) following concentration on SARS-CoV-2 detection was investigated. Eluted concentrated samples from the CP Select concentrator, as discussed in section 2.3, were divided into two parts. AVL lysis buffer with carrier RNA was added into one part at a ratio of 1:1, while the other part was processed without adding the buffer. RNA was extracted from both aliquots using the QIAmp Viral RNA extraction kit, and results were compared with RT-qPCR analysis targeting the N1 gene. The modified CP Select protocol was used in the comparison with the EMF method.

### 2.6 Virus attachment to solid debris

We investigated the possible impact of the centrifugation step on viral recovery. As we separate out solids from the wastewater by centrifugation prior to the filtration with the CP Select concentrator, it is likely that some fraction of viral components may end up settling with the pellet at the centrifugation step (Forés et al., 2021). To quantify the amount of virus settled with the pellet during centrifugation, BCoV spiked wastewater were incubated about an hour at 4°C to attach viruses with the debris properly, then the pellets generated from 80 mL wastewater samples after centrifugation at 10000 g for 10 minutes were extracted using an AllPrep PowerViral DNA/RNA Kit (Qiagen, Cat. No. / ID: 28000-50). Both BCoV recovery and SARS-CoV-2 were quantified from both pellet and supernatant extracts, following the same qPCR protocol used for liquid samples.

### 2.7 Effect of sonication on virus recovery

To assay for increased virus recovery, we tested the effect of sonication, which is known to improve recovery in municipal wastewater treatment plant samples with the PEG and AlCl_3_ precipitation method (Strubbia et al., 2019; Q. Wu & Liu, 2009). A separate set of samples (n=10) were subjected to sonication treatment for 1 minute prior to the centrifugation step, and then processed and quantified as previously described. Equal aliquots of the same set of samples without sonication treatment were processed for comparison. Both BCoV recovery and SARS-CoV-2 (N1 gene) quantification results were considered for this comparison. A summary of sampling sets and sampling volumes used in different experimental setup for the comparison of EMF and CP select method was provided in the supplementary Table S1.

### 2.8 RT-qPCR inhibition

Wastewater is considered as a complex matrix containing a variety of high molecular weight compounds such as humic acids, polysaccharides and proteins that cause interference during RT-qPCR amplification (Schlindwein et al., 2009). This effect may be greater with high concentrations of suspended solids. Though most of the inhibitory substances seem to be removed during the RNA extraction process, residual substances may interfere with the amplification reaction. 10 samples with a 60 mL sample volume were selected to test for the presence of inhibition. RT-qPCR inhibition was assayed by running a VetMAX™ Xeno™ Internal Positive Control - VIC™ Assay (Catalog no. A29767, Applied Biosystems) which has previously been used to test wastewater samples (Greenwald et al., 2021). A known concentration (250 copies/reaction) of VetMAX™ Xeno™ Internal Positive Control (Catalog no-29761, Applied Biosystems) was spiked into RNA extracted from the wastewater and into DNase/RNase free water. VetMAX™ Xeno™ Internal Positive Control - VIC™ Assay was prepared in the same manner as SARS-CoV-2 assay described in section 2.4, only, we added 0.8 uL of premix VetMAX™ Xeno™ - VIC™ Assay instead of N1 primers/probe mix. RT-qPCR was run following the same thermocycling condition as SARS-CoV-2 protocol. All samples were processed together in the same plate to avoid introduction of nuisance variables. The Cq value found in the DNase/RNase water acts as a reference standard for the wastewater sample. If a higher Cq value is measured in wastewater samples compared to the reference Cq value, it is assumed that there is some degree of inhibition due to the composition of the wastewater sample. Typically, a delayed Cq of 2 or greater in wastewater samples relative to the reference Cq value is considered to have RT-qPCR inhibition (Staley et al., 2012; Ahmed et al., 2020b).

### 2.9 Data analysis

All the figures were plotted using Excel 2016 (Microsoft). One-way anova test, t-test and regression analysis were performed using Minitab® 19. P values less than 0.05 were considered statistically significant while greater than 0.05 were considered insignificant or alternative hypotheses are valid. All the RT-qPCR data were analyzed using CFX Maestro™ Software (Biorad).

## 3. Results and discussion

### 3.1 Optimization of CP Select protocol

The automated CP Select concentrator is relatively a new method that has only recently begun to be widely adopted for filtration of wastewater samples. Though there are manufacturer-recommended protocols for concentration of virus from wastewater, which were initially followed, we tested several modifications aimed at improving the performance of the concentration workflow to increase recovery of SARS-CoV-2. Using the manufacturer-recommended protocol we were able to detect SARS-CoV-2 successfully by filtering 100 to 150 mL of wastewater; however, quantification was not as robust as with our established EMF protocol which uses a 40 mL input volume (supplementary Table S2). We had previously observed improved results with EMF filtration upon addition of AVL lysis buffer to the filtered sample. Therefore, we tested the impact of adding an AVL lysis buffer with carrier RNA to the concentrated samples eluted from the CP Select as described in section 2.3. This addition to the manufacturer-recommended protocol significantly improved detection. SARS-CoV-2 was detected in all three replicates from the eluent with added lysis buffer, and not detected in the replicates without the lysis buffer (supplementary Table S3). This optimization step was included in the protocol and when compared with the EMF method, as described in section 3.4, the modified CP Select protocol performed better.

### 3.2 Time comparison of EMF and CP Select concentration methods

A side-by-side comparison of the EMF and the CP Select methods was designed, as shown below in Table 1, to understand how the viral concentration method would impact wastewater sample processing time. There are two components to the processing time - preprocessing and filtration (concentration). Preprocessing consists of sample pH adjustment and MgCl_2_ addition for the EMF method, while centrifugation is used as a preprocessing step for the CP Select protocol. The second step in the protocol is filtration (concentration) itself. We found that for filtration of 40 mL samples, which is the typical input for the EMF protocol in our previous work (Gibas et al. 2021), the CP Select method gave no clear advantage over EMF in filtration time. However, for larger samples of 60 mL and above, the CP Select outperformed the EMF method significantly. For 60 mL samples, the average time to concentration with the CP Select was 9.25 minutes, compared to over 30 minutes for the EMF method. For 100 mL samples, the EMF method could not be used to process most samples, while the CP Select continued to successfully filter samples in under 30 minutes. In addition, when the lab team compared the time required to complete both the preprocessing and the filtration for 20 samples, three hours were required for processing 40 ml using the vacuum manifold EMF approach, while only two hours were required for processing 80 ml when using the CP Select concentrator; this is considering that 6 vacuum manifold stations were available to be used in parallel, and only 4 InnovaPrep stations could be used in parallel. Given this, the CP Select is the practical choice for larger total input volume in routine processing.

### 3.3 Surrogate virus recovery for EMF and CP Select concentration methods

Surrogate virus recovery data is essential to test virus concentration methods as well as process control for the surveillance system especially when RNA of the target organisms cannot be quantified exactly or is difficult to determine. A known concentration of a surrogate virus is spiked into the wastewater before processing and quantified using RT-qPCR following RNA extraction to determine what percentage of the spiked input is recovered from the system, and how much is lost during the sample processing steps. Based on the type of virus concentration method and the RNA extraction process, RNA recovery percentages vary widely. This is often a determining factor for selecting potential virus concentration methods from among different alternatives (LaTurner et al., 2021).

Several different viruses have been used as process controls in WBE studies, including Murine Hepatitis Virus (MHV) (Ahmed, et al., 2020a), Beta Coronavirus OC43 (Pecson et al., 2021; Sherchan et al., 2020) Feline calicivirus (Barril et al., 2021), Human coronavirus (HCoV 229E) (Betancourt et al., 2021; La Rosa et al., 2020), Bovine respiratory syncytial virus (Gonzalez et al., 2020), BCoV (Gonzalez et al., 2020; Jafferali et al., 2021), and Phi 6 (Pecson et al., 2021; Sherchan et al., 2020), MS2 (Rusiñol et al., 2020). Non-enveloped viruses like MS2, when used as a process control, showed higher recovery than enveloped viruses. Enveloped viruses have a lipid layer in the outer membrane making it more susceptible to pH, temperature, and organic solvent (Ye et al., 2016; Polo et al., 2020). We selected BCoV as a process control because it is as an enveloped virus similar to SARS-CoV-2, and belonging to the same Coronaviridae family (LaTurner et al., 2021) as recommended by Pecson et al., (2021) and Sherchan et al., (2020).

For surrogate virus recovery analysis, wastewater samples (n=10) were processed using the same input volume of wastewater (40 mL, 60 mL, and 100 mL) for both the concentration methods side by side. Figure 1 shows the mean BCoV recovery from wastewater concentrated using EMF and the CP Select for different sampling volumes. Both methods showed a wide range of BCoV recovery, due to high variability of sample characteristics such as turbidity. The sample volume also has a role in the variation of BCoV recovery for both concentration methods. The EMF method showed an average BCoV recovery of 17.3% when 40 ml wastewater was filtered which was higher than that for 60 ml and 100 ml sampling volume (Figure 1(b). This is similar to the BCoV recovery rate found by Jafferali et al (2021) and is higher than the reported value of 4.8% by Gonzalez et al (2020); however, this is lower compared with MHV recovery reported in Ahmed et al., (2020). It might be because of the different structure or iso-electric point of MHV compared to BCoV. For the CP Select method, 60 ml sampling size seemed to be optimum based on the BCoV recovery result shown in Figure 1(a). The CP Select method yielded an average of 36.81% BCoV recovery from the 60 ml sampling volume, the highest among all other sampling volumes. Similar results reported in other studies using MS2 and OC43 recovery (Forés et al., 2021; McMinn et al., 2021). In this study, when comparing the BCoV recovery between the two methods under consideration, the CP Select method showed higher recovery than EMF in terms of both median value and average recovery value, as shown in Figure 1, however, it was not statistically significant (P value of 0.12). Forés et al., (2021) also compared two rapid concentration method – CeUF based Centricon plus® 70 and CP Select, and found similar performances in terms of MS2 recovery and SARS-CoV-2 quantification, although higher MHV recovery was reported with the Centricon plus® 70.

**Figure 1:**
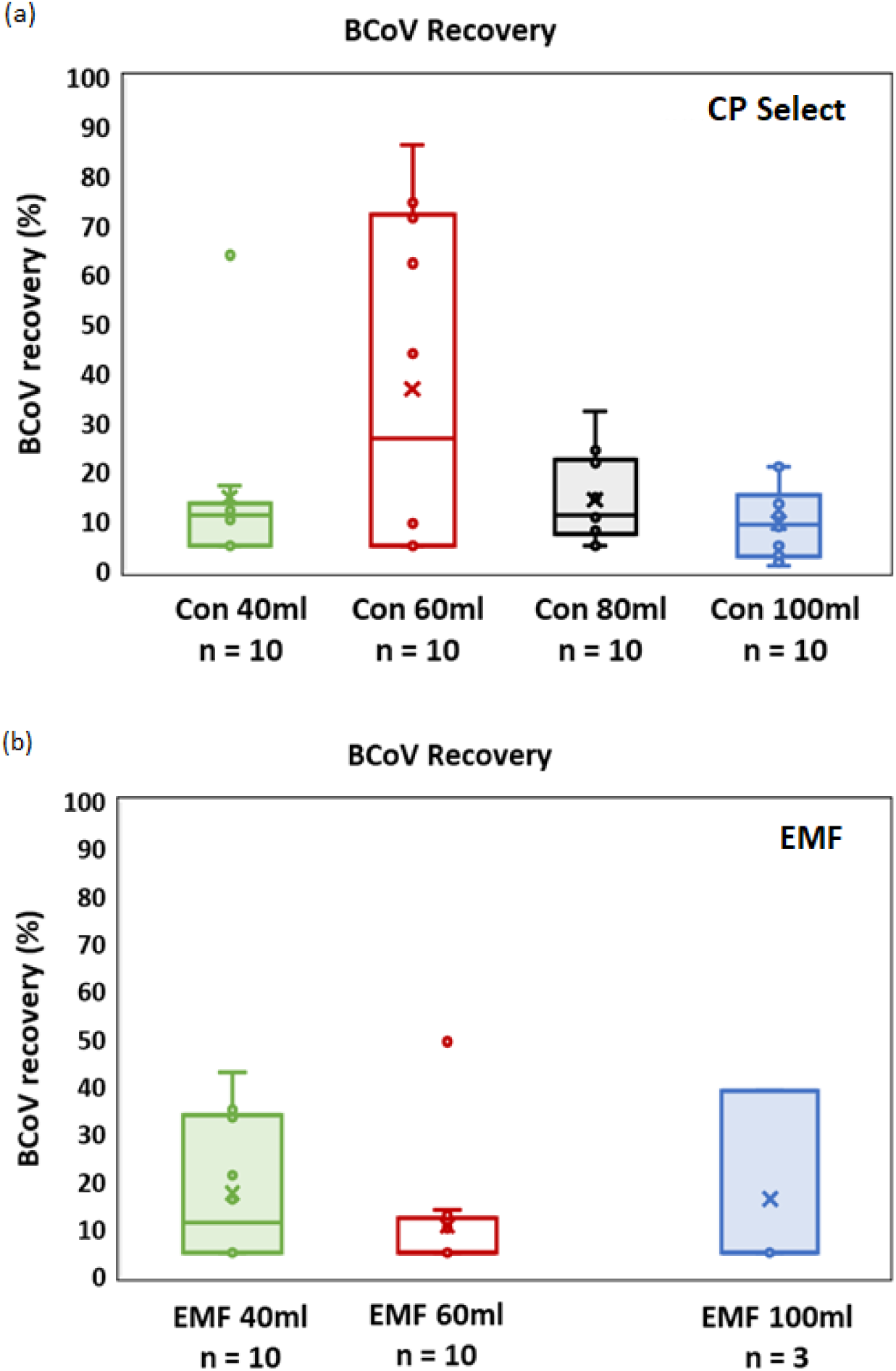
The effect of sampling volume on the BCoV recovery from wastewater sample processed with the CP Select and EMF method. (a) Percentage BCoV recovery for the CP Select method; (b) Percentage of BCoV recovery for the EMF method. The ‘Box’ symbol (□) of the boxplots represents lower (Q1) and upper quartile (Q3) data with median value; ‘Cross’ symbol (×) indicates the average BCoV recovery data. ‘Whiskers’ symbol (工)indicates the data variability outside of the lower and upper quartile with minimum and maximum value.

The effective volume assayed in the RT-qPCR reaction is another way to evaluate the efficiency of concentration methods. Figure 2 shows the comparison of the effective volume assayed in the RT-qPCR reaction depending on the concentration method. The CP Select method allowed the use of up to 5 mL equivalent wastewater per reaction, with a minimum of 1.33 mL, while the range of effective volume for the EMF method was 0.66 - 1.67 mL. Similarly, use of the CP Select also resulted in higher concentration factors ranging 160-600× in comparison to 40-100× with the EMF method.

**Figure 2:**
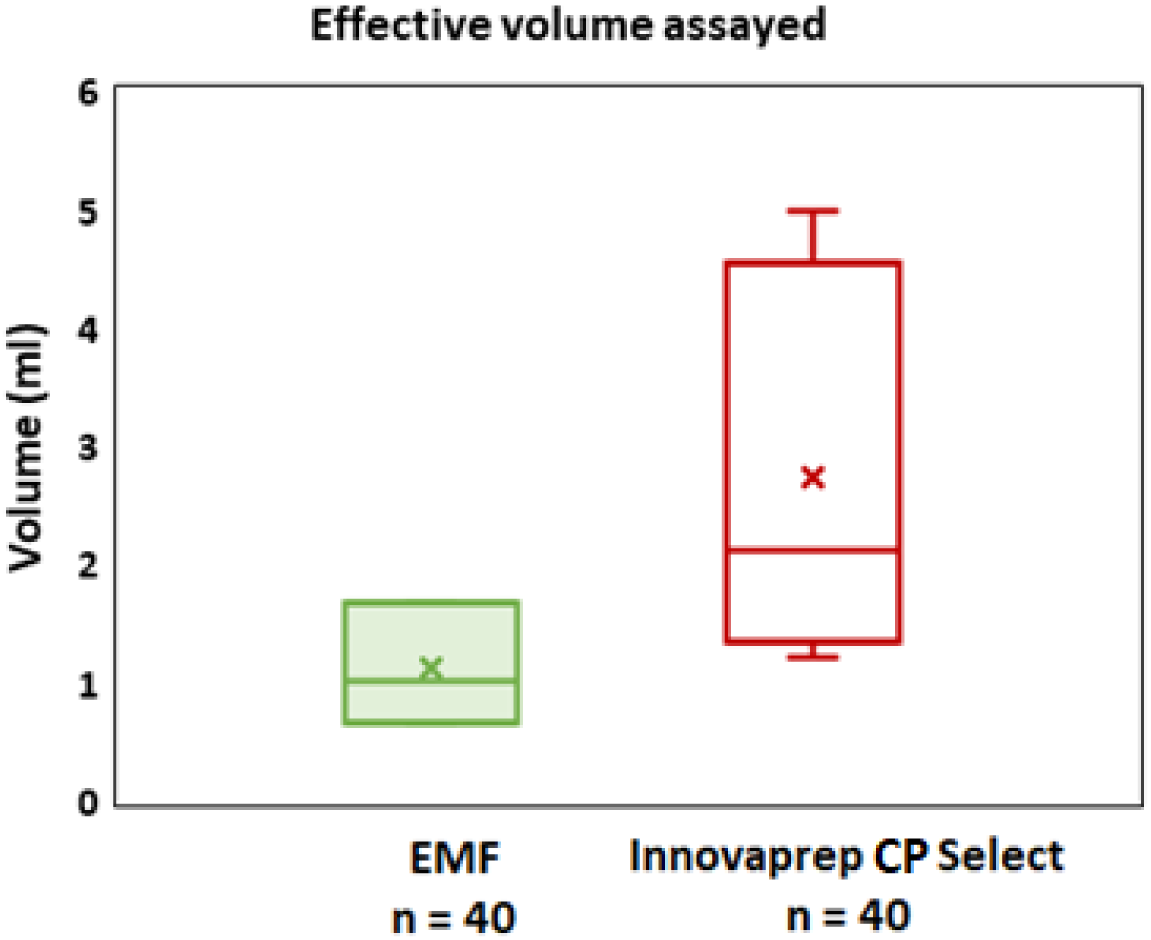
Effective volume assayed to RT- qPCR reaction concentrated with EMF and CP Select methods. The ‘Box’ symbol (□) represents lower (Q1) and upper quartile (Q3) data with median value; ‘Cross’ symbol (×) indicates the average effective volume used in the RT-qPCR reaction. ‘Whiskers’ symbol (工)indicates the data variability outside of the lower and upper quartile with minimum and maximum effective volume.

### 3.4 Performance comparison based on SARS-CoV-2 detection and quantification

Surrogate virus recovery may not be the best indicator of natural SARS-CoV-2 detection in real wastewater samples (Forés et al., 2021), As such, we also compared performance between the CP Select and EMF concentration method in terms of SARS-CoV-2 detection and quantification using the same samples from the previous sections. Table 2 compares results from both methods based on 40 mL and 60 mL input volumes. The EMF approach could not reliably be used to process 100 mL samples as listed in Table 1, and so a direct comparison of outcomes from the two methods for that sample volume was not possible. Both methods successfully detected naturally occurring SARS-CoV-2 virus from the wastewater, however, the CP Select method was more sensitive as shown in Table 2.

**Table 2:**
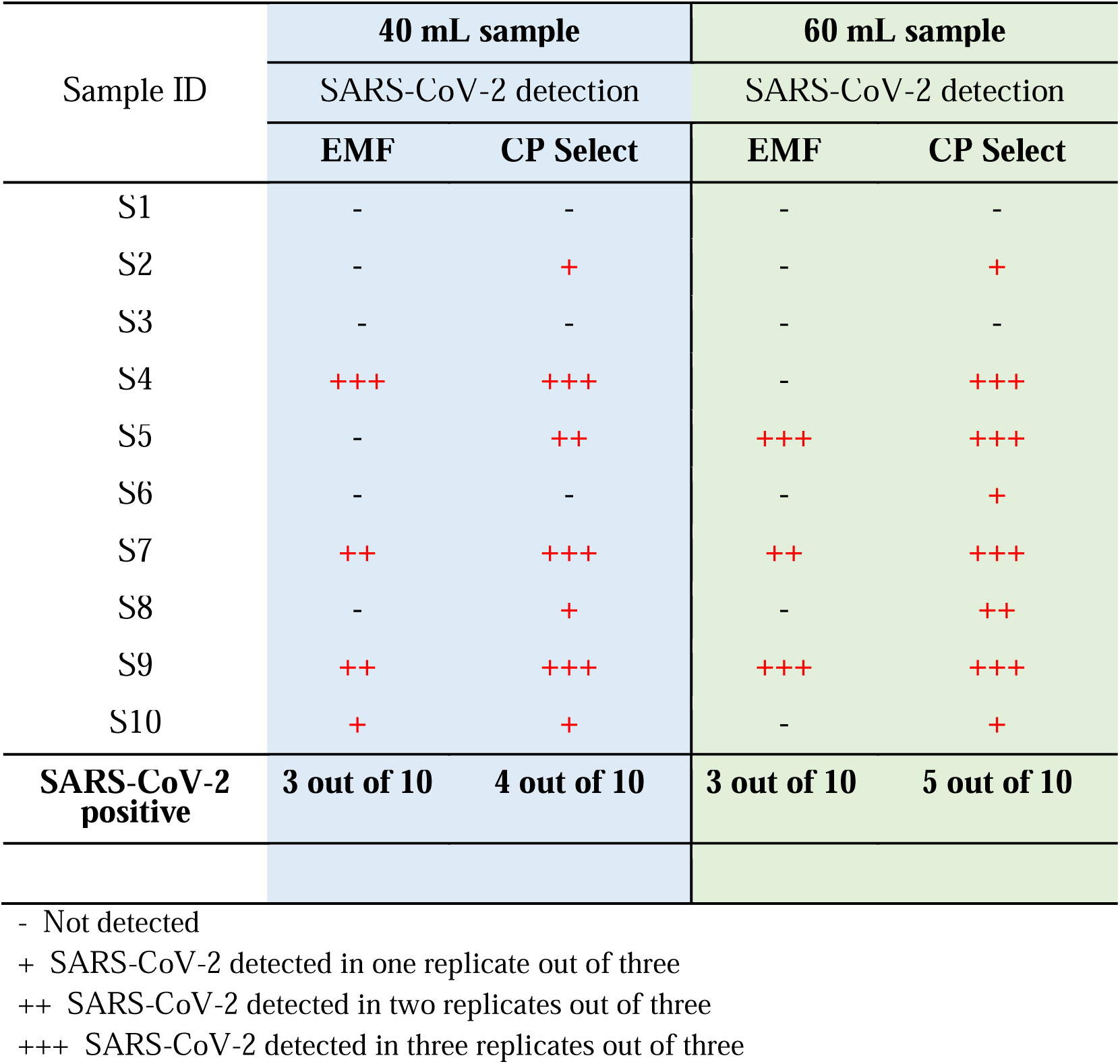
SARS-CoV-2 detection from wastewater sample concentrated by EMF and CP Select methods.

For the 40 mL sampling volume, three samples were detected as SARS-CoV-2 positive using the EMF protocol while four samples were positive when the CP Select protocol was used. When a 60 mL sample volume was used as input, no additional positives were detected using the EMF protocol, but two additional samples were detected as positive with the CP Select protocol. Detection was also more robust following CP Select processing with the larger sample; all three qPCR replicates were positive in more samples in contrast to EMF-processed samples not showing detectable amplification in all replicates (supplementary Table S4). Also, the variation among Cq values for each sample using the CP Select method was lower.

The LoD for the CP Select assay workflow was in the range of 1.5⍰10^3^ to 3.75⍰10^3^ copies/L for 100 mL to 40 mL wastewater samples processed, respectively, while it was 3.0⍰10^3^ to 7.5⍰10^3^ copies/L for the EMF method; twice the LoD of the CP Select method. Figure 3(a) and 3(b) which show the variability in viral copy number detected from the same set of samples using the EMF and CP Select workflows. When a 40 mL sample was processed using the EMF, SARS-CoV-2 quantification ranged from 10^4^ - 4.2 10^5^ genome copies/L while it was 1.5 10^3^ - 9.3 10^4^ genome copies/L using the CP Select method. The lower end of the quantification range of the latter method is due to the samples that did not amplify and could be considered non-detected with the EMF method. However, no significant differences were observed between these two methods in SARS-CoV-2 quantification for high titer wastewater samples (P = 0.51). This described trend was also observed for the 60 mL data set. Other studies also reported similar range of SARS-CoV-2 concentration both in the university resident hall’s wastewater and WWTP’s influent using the CP Select method. For example, A range of 2.4⍰10^4^ - 4⍰10^4^ copies/L of SARS-CoV-2 gene was reported in the residence hall wastewater using the CP Select method during the surveillance at the University of Windsor (Corchis-Scott et al., 2021) while it was in the range from 10^3^ - 2⍰10^5^ copies/L for WWTP’s influent samples (Forés et al., 2021; Lynch et al., 2021). Although these data are mostly similar to the concentration found in this study, however, there is highly likely to have a different concentration of SARS-CoV-2 in wastewater as it mostly depends on the density of the COVID-19 infected people staying in those area during the sampling time, demographic location, pattern of the sewerage system, and wastewater characteristics (Barua et al., 2021).

**Figure 3.**
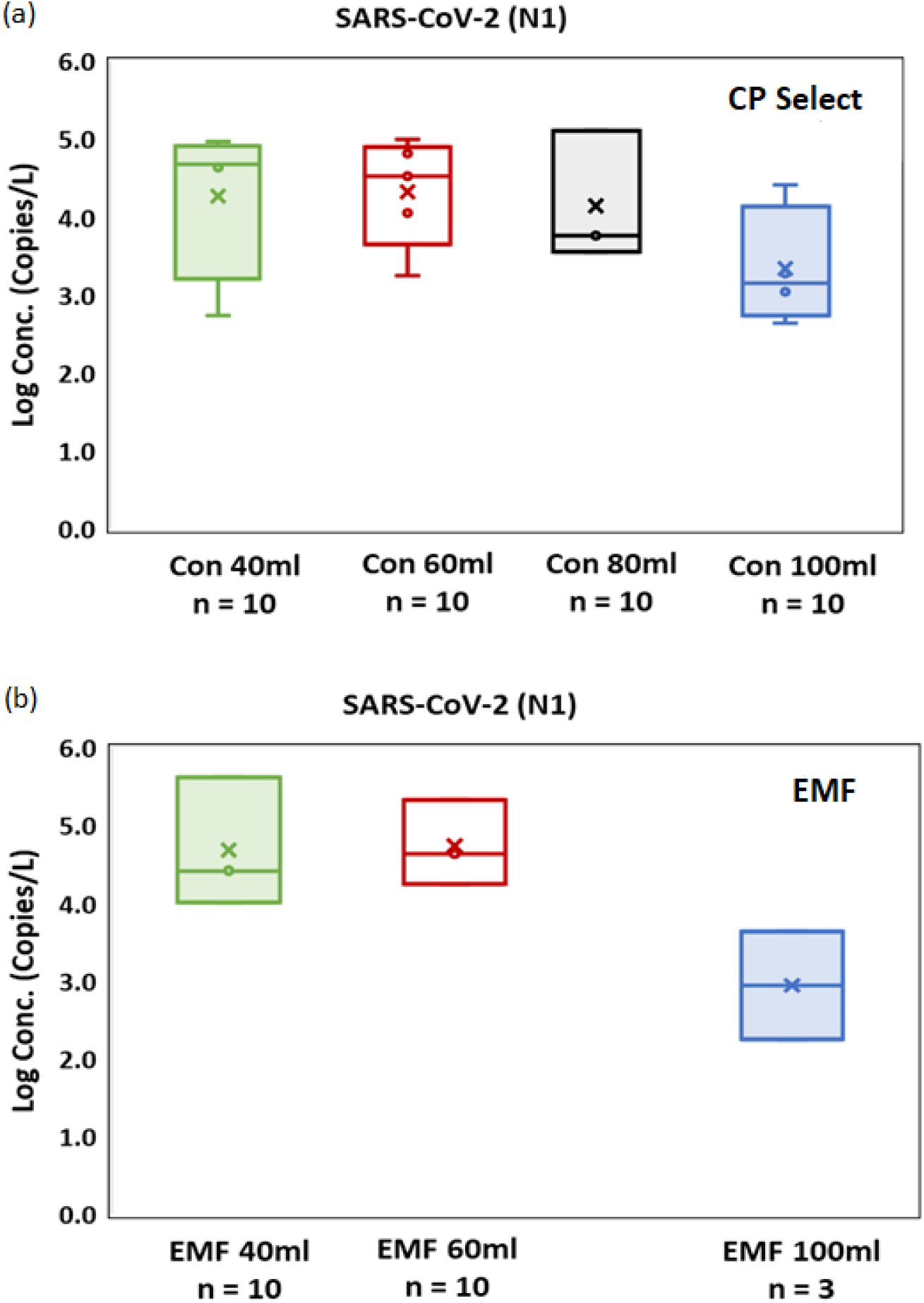
Effect of sample volume size on the performance of CP Select concentrator and EMF in terms of SARS-CoV-2 quantification. (a) SARS-CoV-2 quantification from concentrated samples using Innovaprep CP Select concentrator; (b) SARS-CoV-2 quantification from concentrated samples using EMF. The ‘Box’ symbol (□) represents lower (Q1) and upper quartile (Q3) data with median value; ‘Cross’ symbol (×) indicates the average SARS-CoV-2 quantification data. ‘Whiskers’ symbol (工)indicates the data variability outside of the lower and upper quartile with minimum and maximum Log transformed SARS-CoV-2 concentration.

SARS-CoV-2 detection and quantification performance was further evaluated for the CP select method using a larger input volume of wastewater, on the assumption that concentrating more copies of the virus, would allow for better quantification of low viral titer samples. A separate set of samples (n=20) were then processed using the two concentration methods side by side, followed by RNA extraction, and quantification. 100-150 mL wastewater was filtered through the CP Select concentrator, while 40 mL (the volume routinely used in our surveillance protocol) was filtered through the EMF filter. Out of 20 wastewater samples, SARS-CoV-2 was detected in 8 samples processed with the EMF filtration, while 12 samples were positive when processed using the CP Select (Figure 4). By concentrating viruses from a larger volume of wastewater, the CP Select method resulted in more sensitive detection. Five samples reported negative using the routinely followed EMF method were detected as SARS-CoV-2 positive when processed with the CP Select method, while in only one case did EMF filtration detect a positive where the CP Select did not. The SARS-CoV-2 was detected in the additional CP Select derived samples had higher Cq values (i.e at lower viral copy numbers) which indicated that the workflow using the CP Select concentration step is capable of capturing viruses from low-titer wastewater samples that may be missed using the EMF method.

**Figure 4:**
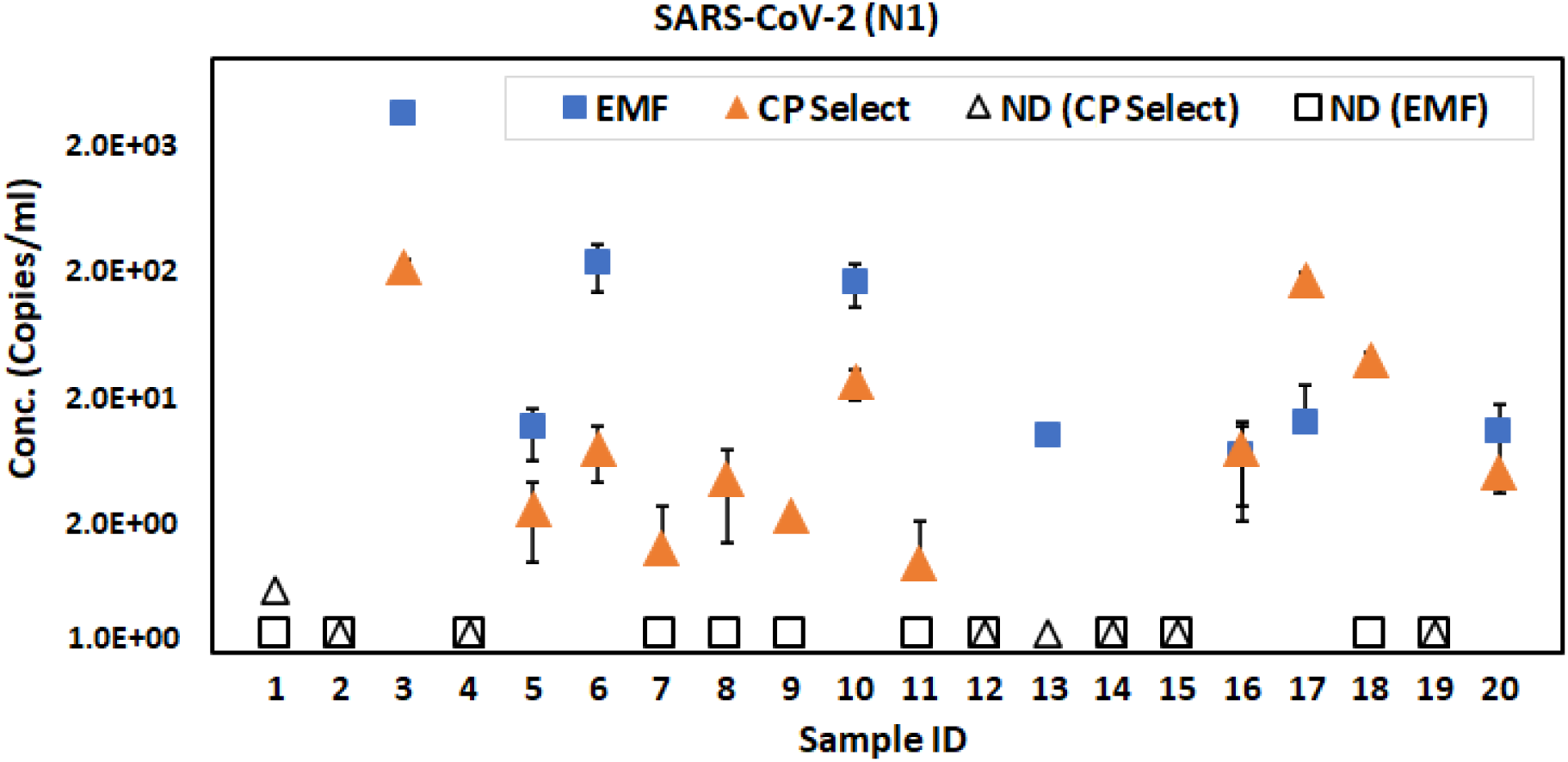
Detection of SARS-CoV-2 from wastewater concentrated using the EMF and the CP Select method. Out of 20 samples, the EMF method detected natural SARS-CoV-2 in 8 samples while CP Select detected in 12 samples. Solid **blue square** boxes indicate SARS-CoV-2 positive samples processed with the EMF method and **orange triangles** indicate SARS-CoV-2 positives with the CP Select method. **Open symbols** indicate the non-detected samples with the corresponding method. Error bars indicate the standard deviation among replicates. LoD is 1.37 copies/mL for the CP Select method (150 mL sample size) while 7.5 copies/mL for the EMF method (40 mL sample size)

Overall, the CP Select method is more sensitive than EMF method as the higher number of positive samples obtained by this method than obtained by the EMF method. The CP Select method is beneficial in situations where detection sensitivity and quick data reporting is important. The tradeoff for this method is the cost effectiveness where the CP Select protocol doubles the material and the reagent cost per sample as compared with the EMF method. However, the ease of operation of the CP Select also reduces the number of lab workers and time needed to process the samples.

### 3.5 Virus attachment to solid debris

To determine whether a significant amount of virus remained in the pellets following centrifugation step, we quantified recovery of BCoV and natural SARS-CoV-2 from both the pellet and the supernatant of centrifuged samples (Figure 5). A significantly smaller fraction of BCoV was recovered from the pellet than from the supernatant, with a P-value of 0.015 (P < 0.05). However, SARS-CoV-2 behaved differently from BCoV in centrifugation, with similar recovery fractions in the supernatant and the pellet (P value of 0.857). This difference may be due to the viral structure itself; the structure of the spike protein may result in SARS-CoV-2 attaching more strongly to a solid surface compared to BCoV. Ai et al., (2021) reported a similar trend with 0.2% BCoV recovery from the pellets while the SARS-CoV-2 recovery was found to be 10%. Similar to the SARS-CoV-2 partitioning results found in our analysis Forés et al., (2021) reported about 23% SARS-CoV-2 recovery from the pellet, and Ye et al., (2016) observed about 24% MHV partitioning to the solid. However, another study reported no significant difference in SARS-CoV-2 quantification results due to separating solids from the liquid (Pecson et al., 2021), although the pellet material was not directly assayed. The variation in recoveries observed in different studies may be due to the variability in the wastewater matrix at different location or collection sites, or due to the differences in the methodological approaches.

**Figure 5:**
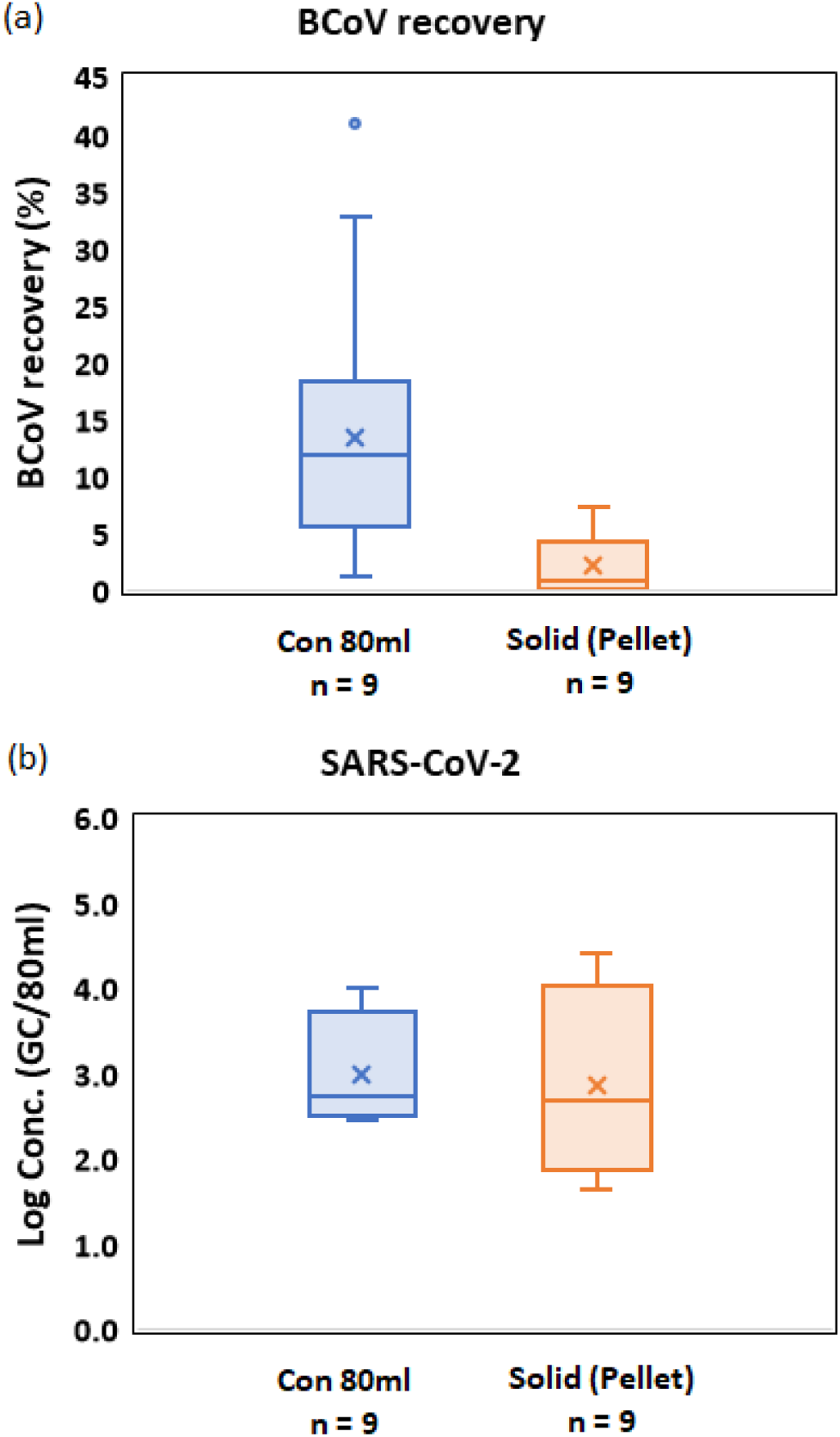
Fraction of viral material partitioned to the supernatant and solid debris fraction for CP Select processed samples, which are centrifuged prior to concentration to remove debris. (a) percentage of BCoV recovery and (b) SARS-CoV-2 quantification. The ‘Box’ symbol (□) of the boxplots represents lower (Q1) and upper quartile (Q3) data with median value; ‘Cross’ symbol (×) indicates the average value of the data set. ‘Whiskers’ symbol (工)indicates the data variability outside of the lower and upper quartile with minimum and maximum value.

### 3.6 Effect of sonication on virus recovery

In the previous section, we observed that a fraction of viral material is adsorbed by suspended solids and settled with the pellet during centrifugation step. To counter this effect, we tested the impact of a very short sonication step (1 minute) prior to centrifugation of wastewater samples. The sonication step acts to disrupt the attachment of viral material to solids but was kept short to minimize damage to the viral RNA itself. Sonication treatment has been previously shown to increase viral recovery by causing desorption of viral particles from organic substances and release of viral particles from host cells (Corpuz et al., 2020; Strubbia et al., 2019).

Results of the sonication experiment are shown in Table 3. BCoV recovery improved for most samples after addition of the sonication treatment. Average recovery increased from 3.85% to 23.74%. Due to the variability of material collected during our ongoing sampling operation and available for testing the group of samples for this analysis were very turbid (Table 3) compared to some of the samples used previously (Table 1), and initial BCoV recovery from these samples was somewhat lower than typical. Along with improved BCoV recovery from a majority of samples, SARS-CoV-2 detection also improved with sonication treatment, with Cq values being lower in many instances, and detection of the virus in samples which had previously appeared to be negative (Table 3). The sonication step may partly solve a problem common to all ultrafiltration-based concentration methods, in which some part of the virus is lost with the pellet during centrifugation. We subsequently adopted the sonication step as part of our standard CP Select virus concentration protocol used for the routine SARS-CoV-2 wastewater-based monitoring at UNC Charlotte.

**Table 3:** The effect of sonication treatment on BCoV recovery and SARS-CoV-2 detection.

| Sample ID | pH | Turbidity (NTU) | BCoV recovery |  |  |  | SARS-CoV-2 |  |
| --- | --- | --- | --- | --- | --- | --- | --- | --- |
|  |  |  | Without Sonication |  | With Sonication |  | Without Sonication | With Sonication |
|  |  |  | Avg Cq | Recovery (%) | Avg Cq | Recovery (%) | Avg Cq | Avg Cq |
| S31 | 7.5 | 46.5 | 33.7 | 3.2 | 29.8 | 50.4 | 36.2* | 35.0 |
| S32 | 7.5 | >1000 | 35.1 | 1.2 | ND | - | 35.2* | ND |
| S33 | 8 | 390 | 33.5 | 3.8 | 33.5 | 3.8 | ND | 34.9 |
| S34 | 7.5 | 338 | 35.1 | 1.3 | 34.5 | 1.8 | ND | 40.0 |
| S35 | 7.5 | >1000 | 35.2 | 1.1 | 32.6 | 7.1 | 35.5 | 37.5 |
| S36 | 8.5 | 38.2 | 34.2 | 2.3 | 30.0 | 59.8 | 34.7 | 32.3 |
| S37 | 8 | >1000 | 33.0 | 5.2 | 31.6 | 14.5 | 36.5 | 35.7 |
| S38 | 8 | 58.2 | 31.7 | 12.9 | 30.6 | 28.8 | 35.5 | 35.9 |
| S39 | 8 | 978 | ND | - | ND | - | ND | ND |

### 3.7 qPCR inhibition

RT-qPCR detection of the VetMAX™ Xeno™ Internal Positive Control spiked into the extracted RNA is shown in Figure 6. An average Cq of 8 NTC replicates was used as the reference point (Cq = 32.62). Most samples did not appear to be affected by inhibitors in the RT-qPCR step using either protocol, as nearly all Cq values fall within 2 Cq of the reference line. One sample processed with the CP Select did show a delayed Cq, which was not replicated when the sample was processed using EMF, but overall, the difference between the two methods did not meet a threshold for statistical significance when all values were compared. Cq values for all other samples processed with both methods were within the 1 Cq variation from the reference value. This suggests there is no consistent and significant inhibition to RT-qPCR amplification for extracted RNA from samples processed with either of the two filtration methods.

**Figure 6:**
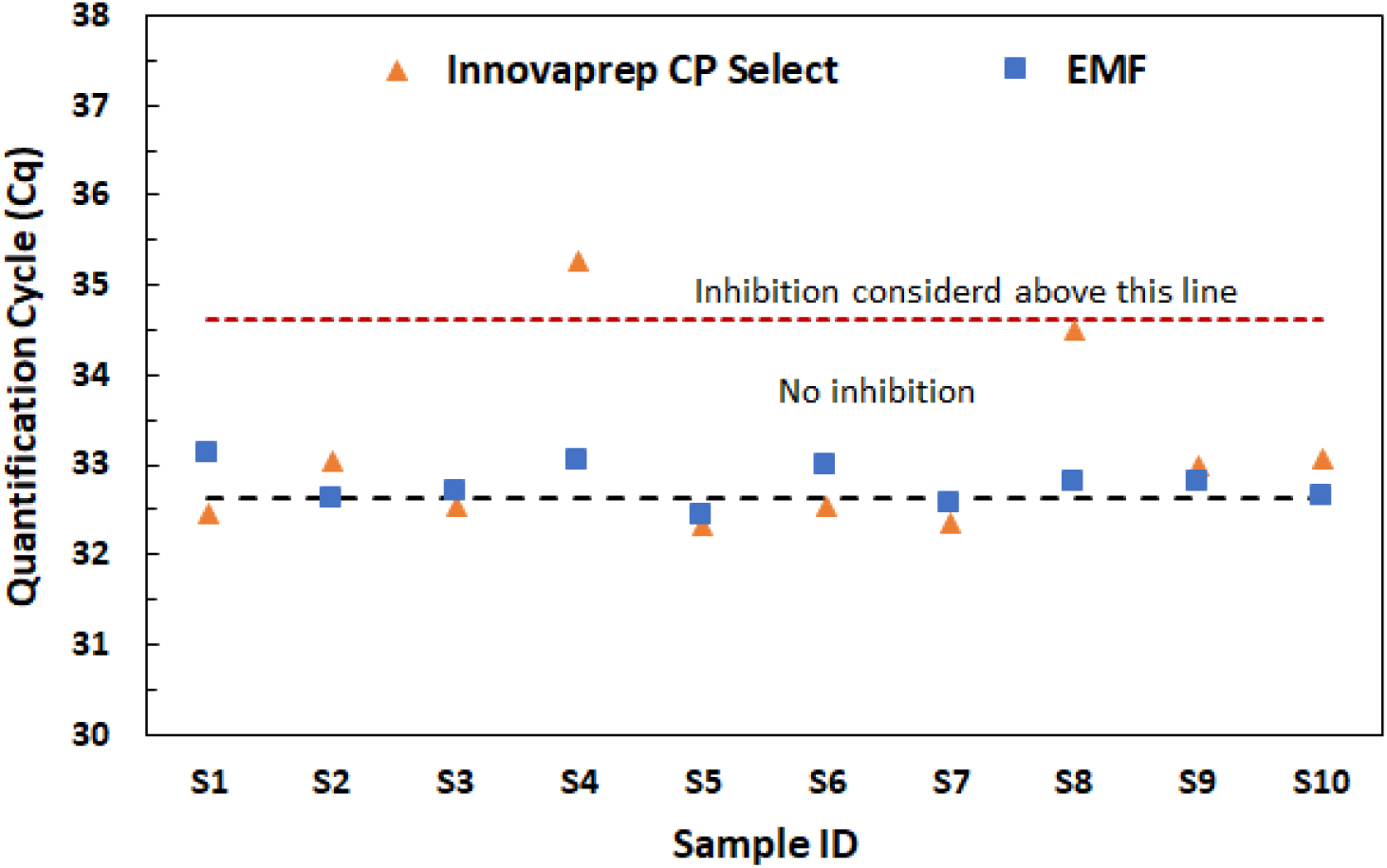
RT-qPCR inhibition test comparing results for samples concentrated with CP Select and with the EMF method. Across all samples, differences in Cq did not rise to the level of statistical significance.

## 4. Conclusion

We have developed an optimized protocol for use of the InnovaPrep CP Select concentrator, in a routine building wastewater surveillance program on a university campus. The CP Select method resulted in a BCoV recovery rate of approximately 37%, which is higher than BCoV recovery from samples processed using an EMF protocol. The CP Select is capable of processing up to 150 mL of wastewater within 30 minutes, while the EMF method fails at larger volumes and operates optimally with 40 mL input. This allows for a higher effective volume of wastewater to be assayed with the CP Select relative to EMF, which in turn results in increased sensitivity for detection and quantification of SARS-CoV-2 from wastewater. Overall, the processing time for handling a typical day’s collected samples in a surveillance scenario was decreased by 33% (from 3 hours to 2 hours). We found that use of a lysis buffer (AVL) significantly improved the performance of the InnovaPrep manufacturer recommended protocol for wastewater and have introduced that modification to our routine work. One observation in use of an ultrafiltration-based protocol was that viral material may be lost with the pellet in the required centrifugation step, however, in combination with a brief sonication treatment, we were able to achieve higher recovery fractions. We did not observe significant differences in qPCR inhibition when the CP Select protocol was used, relative to the EMF protocol. In general, the CP Select concentrator is advantageous for concentrating low viral titer wastewater samples, especially when rapid data reporting is necessary, and use of this protocol can also improve recovery and detection sensitivity.

## Data Availability

Data is provided in the supplementary file and also available from the authors upon request.

## Acknowledgements

We would like to thank Richard Tankersley, Vice Chancellor for Research and Economic Development and his team for supporting this project, and Rachel Noble, Professor at UNC-IMS for sharing knowledge regarding wastewater sample processing protocols that was leveraged in this project, Greg Cole and his team in Facilities Management for plumbing support and rest of members of the UNC Charlotte wastewater monitoring group including Visva B. Barua, Neha Mittal, Lauren R. Brazell, Keshawn Hinton, Isaiah Young.

---

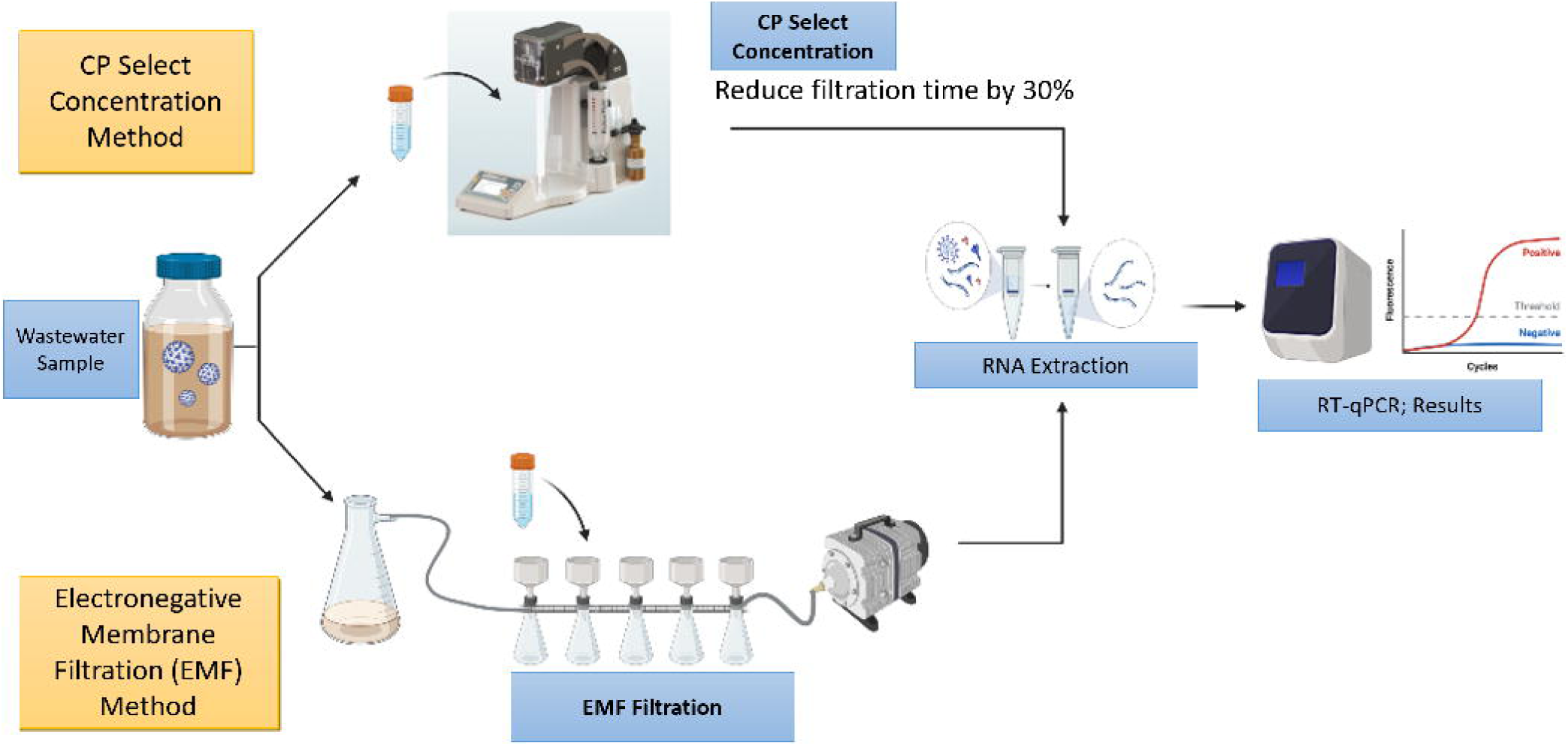

