## Supplementary Files for "Performance evaluation of virus concentration methods for implementing SARS-CoV-2 wastewater based epidemiology emphasizing quick data turnaround"

**Supplementary Table S1:** Summary of sampling set and sampling volume used in different experimental setup for the comparison of EMF and CP select methods.

| Sample ID | No. of samples | Volume  (ml) | Objectives and sections covered |
| --- | --- | --- | --- |
| S1 to S10 | 10 | 40 – 100 | Time comparison of EMF and CP Select concentration methods (section 3.2) |
|  |  | 40 - 100 | Surrogate virus recovery for EMF and CP Select concentration methods (section 3.3) |
|  |  | 40 - 100 | Performance comparison based on SARS-CoV-2 detection and quantification (section 3.4) |
|  |  | 80 | Virus attachment to solid debris (section 3.5) |
|  |  | 60 | qPCR inhibition (section 3.7) |
| 1 to 20 | 20 | 40*/150* | Impact of higher sample volume on CP Select in terms of SARS-CoV-2 detection and quantification (section 3.4, Figure 4) |
| 31 to 39 | 9 | 60 | Effect of sonication on virus recovery (section 3.6) |

*40 ml used in EMF while 125-150 ml used in the CP Select method.

**Supplementary Table S2**: Comparison between two concentration methods. In this analysis, 40 mL wastewater was filtered using EMF and 125 to 150 mL processed with the CP Select Concentrator. The AVL lysis buffer was not added to the concentrated sample from the CP Select protocol. Water quality includes in the Table.

| **Sample name** | **pH** | **Turbidity (NTU)** | **EMF** | | | **CP Select** | | |
| --- | --- | --- | --- | --- | --- | --- | --- | --- |
|  |  |  | Cq | Mean Cq | Copies/L | Cq | Mean Cq | Copies/L |
| Greek-3_10.16.20 | 7.0 | 488 | ND | 35.82 | **1.46E+04** | ND | 34.87 | **2.31E+03** |
|  |  |  | 35.74 |  |  | 35.25 |  |  |
|  |  |  | 35.9 |  |  | 34.48 |  |  |
| Holshouser_10.16.20 | 8.0 | 45.2 | 35.65 | 35.80 | **1.48E+04** | ND |  |  |
|  |  |  | 35.79 |  |  | 36.23 |  |  |
|  |  |  | 35.95 |  |  | ND |  |  |
| Greek-3_10.14.20 | - | - | ND | 35.16 | **2.35E+04** | 35.15 | 34.85 | **2.31E+03** |
|  |  |  | 34.89 |  |  | 34.81 |  |  |
|  |  |  | 35.42 |  |  | 34.59 |  |  |
| Holshouser_10.14.20 | 8.5 | - | Negative |  |  | Negative |  |  |
| Greek-3_10.12.20 | - |  | 34.42 | 35.9 | **3.31E+04** | 38.40 | 38.44* | **1.14E+03** |
|  |  | - | 36.77 |  |  | ND |  |  |
|  |  |  | 36.51 |  |  | 38.48 |  |  |
| Holshouser_10.12.20 | - | - | Negative |  |  | Negative |  |  |
| Lynch_10.16.20 | 7.5 | 183 | Negative |  |  | Negative |  |  |
| Scott_10.16.20 | 8.5 | 200 | Negative |  |  | Negative |  |  |
| Sanford_10.16.20 | 8.5 | 87.7 | ND |  |  | Negative |  |  |
|  |  |  | ND |  |  |  |  |  |
|  |  |  | 36.22 |  |  |  |  |  |
| Hunt_10.16.20 | - | - | Negative |  |  | Negative |  |  |

* 50 ml wastewater was concentrated to 250 uL in the concentrator. Red Color indicates out of the LoD

**Supplementary Table S3:** The effect of addition of lysis buffer to the concentrated eluted sample from the CP Select method.

| Sample ID | With of AVL Lysis buffer | | | | Without AVL lysis buffer | | | |
| --- | --- | --- | --- | --- | --- | --- | --- | --- |
|  | Rep.#1 | Rep.#2 | Rep.#3 | Mean Cq | Rep.#1 | Rep.#2 | Rep.#3 | Mean Cq |
| Hawthorn | ND | ND | ND | - | ND | 40.24 | 36.65 | **38.45** |
| Holshouser | 35.47 | 37.39 | 36.31 | **36.39** | ND | ND | 36.36 | - |
| Belk_West | 32.87 | 32.58 | 33.39 | **32.95** | ND | 39.19 | ND | - |

Red Color indicates out of LoD.

**Supplementary Table S4:**  Effect of sample volume size on the performance of EMF and CP Select methods in terms of SARS-CoV-2 quantification.

| **EMF filtration (40 mL)** | | | | | |  | **CP Select (40 mL)** | | | | |
| --- | --- | --- | --- | --- | --- | --- | --- | --- | --- | --- | --- |
| **I.D.** | **Cq Value** | | | |  |  | **Cq Value** | | | |  |
|  | Rep. #1 | Rep. #2 | Repl. #3 | **Mean Cq** | **SD** |  | Rep. #1 | Rep. #2 | Repl. #3 | **Mean Cq** | **SD** |
| S1 | - | - | - | - |  |  | - | - | - | - |  |
| S2 | - | - | - | - |  |  | 36.14 | - | - | **-** |  |
| S3 | - | - | - | - |  |  | - | - | - | - |  |
| S4 | 31.13 | 31.16 | 31.12 | **31.14** | **0.02** |  | 31.67 | 32.41 | 32.78 | **32.28** | **0.56** |
| S5 | - | - | - |  |  |  | 39.10 | 37.04 | - | **38.07** | **1.45** |
| S6 | - | - | - | - |  |  | - | - | - | - |  |
| S7 | 34.60 | - | 35.40 | **35** | **0.57** |  | 32.90 | 33.15 | 33.54 | **33.2** | **0.32** |
| S8 | - | - | - | - |  |  | 37.09 | - | - | - |  |
| S9 | 36.62 | - | 36.02 | **36.32** | **0.42** |  | 33.22 | 33.14 | 34.05 | **33.47** | **0.50** |
| S10 | 37 | - | - | **-** |  |  | 37.46 | - | - | **-** |  |
| **EMF filtration (60 mL)** | | | | | |  | **CP Select (60 mL)** | | | | |
| **I.D.** | **Cq Value** | | | |  |  | **Cq Value** | | | |  |
|  | Rep. #1 | Rep. #2 | Repl. #3 | **Mean Cq** | **SD** |  | Rep. #1 | Rep. #2 | Repl. #3 | **Mean Cq** | **SD** |
| S1 | - | - | - | - |  |  | - | - | - | - |  |
| S2 | - | - | - | - |  |  | 37.12 | - | - | - |  |
| S3 | - | - | - | - |  |  | - | - | - | - |  |
| S4 | - | - | - | - |  |  | 32.42 | 32.61 | 32.63 | **32.55** | **0.11** |
| S5 | 36.31 | 41.24 | 41.24 | **39.60** | **2.85** |  | 34.70 | 35.02 | 35.54 | **35.08** | **0.42** |
| S6 | - | - | - | - |  |  | 37.23 | - | - | - |  |
| S7 | 34.60 | - | 35.40 | **35** | **0.57** |  | 31.75 | 32.35 | 31.90 | **32.00** | **0.31** |
| S8 | - | - | - | - |  |  | 37.33 | - | 37.16 | **37.24** | **0.12** |
| S9 | 31.75 | 31.17 | 31.66 | **31.53** | **0.31** |  | 32.97 | 32.80 | 33.60 | **33.12** | **0.42** |
| S10 | - | - | - | - |  |  | 37.04 | - | - | **-** |  |
| **EMF filtration (100 mL)** | | | | | |  | **CP Select (100 mL)** | | | | |
| **I.D.** | **Cq Value** | | | |  |  | **Cq Value** | | | |  |
|  | Rep. #1 | Rep. #2 | Repl. #3 | **Mean Cq** | **SD** |  | Rep. #1 | Rep. #2 | Repl. #3 | **Mean Cq** | **SD** |
| S1 |  |  |  |  |  |  | - | - | - | - |  |
| S2 |  |  |  |  |  |  | - | - | - | - |  |
| S3 | - | - | - |  |  |  | - | - | - | - |  |
| S4 |  |  |  |  |  |  | 32.53 | 33.06 | 32.71 | **32.77** | **0.27** |
| S5 | 38.48 | 34.00 | ND | 36.24 | 3.17 |  | 41.79 | 35.41 | - | **38.60** | **4.51** |
| S6 | 38.89 | 42.44 | ND | 40.66 | 2.51 |  | - | - | - | - |  |
| S7 |  |  |  |  |  |  | 37.26 | 37.87 | 36.40 | **37.18** | **0.73** |
| S8 |  |  |  |  |  |  | - | - | - | - |  |
| S9 |  |  |  |  |  |  | 37.23 | 37.17 | 35.66 | **36.69** | **0.89** |
| S10 |  |  |  |  |  |  | - | - | 36.83 | - |  |

Grey color column indicates those samples did not pass through the EMF filter during processing.

**Supplementary Table S5:** Primers and probe sequences used in this study.

| Assay | Primer/Probe | Sequences | References |
| --- | --- | --- | --- |
| CDC N1 | Forward | GACCCCAAAATCAGCGAAAT | Lu et al., 2020 |
|  | Reverse | TCTGGTTACTGCCAGTTGAATCTG |  |
|  | Probe | FAM-ACCCCGCAT/ZEN/TACGTTTGGTGGACC-3IABkFQ |  |
| BCoV | Forward | CTGGAAGTTGGTGGAGTT | Decaro et al., 2008 |
|  | Reverse | ATTATCGGCCTAACATACATC |  |
|  | Probe | FAM-CCTTCATATCTATACACATCAAGTTGTT-BHQ1 |  |

(a)

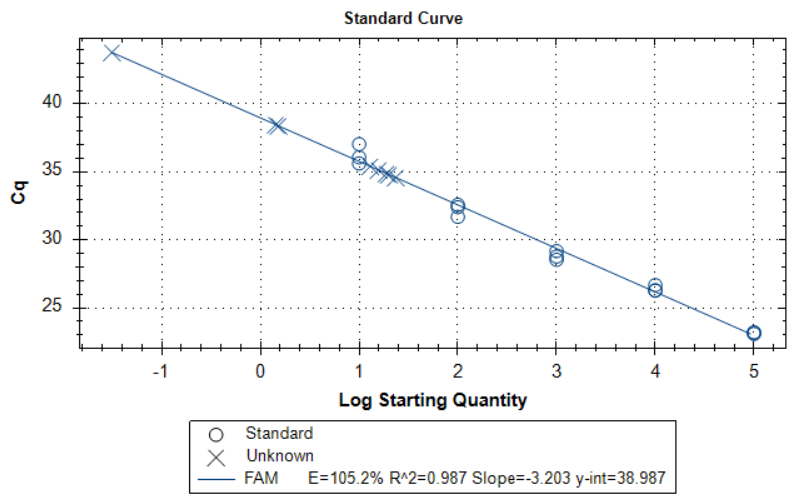

(b)

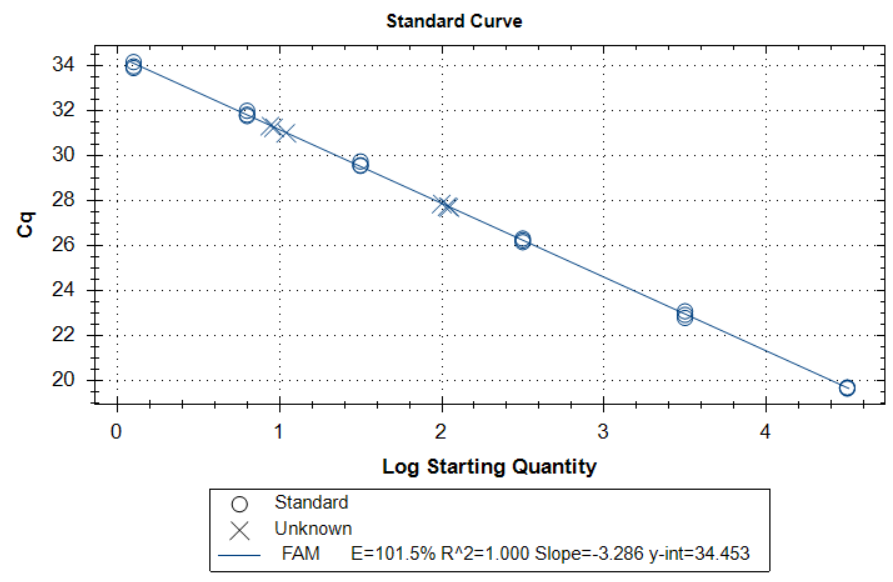

**Supplementary Figure S1**: Standard curve used in this study: (a) SARS-CoV-2 (N1) (b) BCoV

**Supplementary Table S6:** Water quality of the wastewater samples used in the Fig.4

| **Sample ID** | **pH** | **Turbidity (NTU)** |
| --- | --- | --- |
| 1 | 8 | 65.1 |
| 2 | 7.5 | 77.5 |
| 3 | 7.5 | 46.4 |
| 4 | 7.5 | 59.6 |
| 5 | 8.0 | 60.9 |
| 6 | 8.5 | 342 |
| 7 | - | - |
| 8 | 7.5 | 854 |
| 9 | 7.5 | >1000 |
| 10 | 7.5 | 61.5 |
| 11 | 7.0 | 43.5 |
| 12 | 8.0 | 25.4 |
| 13 | 8.5 | 131 |
| 14 | 7.0 | 14.7 |
| 15 | 8.0 | 739 |
| 16 | 9.0 | 265 |
| 17 | 7.5 | 86 |
| 18 | 7.0 | >1000 |
| 19 | 7.0 | >1000 |
| 20 | 7.5 | 64.7 |
